# *THRB* splice site variants lead to exon 4 skipping and TRβ1 gain-of-function syndrome

**DOI:** 10.64898/2026.04.15.26349265

**Authors:** Georg Sebastian Hönes, Xiao-Hui Liao, Elisa Annabelle Mahler, Philipp Herrmann, Anja Eckstein, Dagmar Führer, Jesus Moreno Castillo, John Chiang, Andrea Louise Vincent, Roy E. Weiss, Alexandra Mihaela Dumitrescu, Samuel Refetoff, Lars Christian Moeller

**Author notes:** **Correspondence** Prof. Dr. Lars Moeller, Department of Endocrinology, Diabetes and Metabolism, University Hospital Essen, University of Duisburg-Essen, Hufelandstr. 55, 45147 Essen, Germany.

## Abstract

**Background:** Heterozygous c.283+1G>A and c.283G>A variants in the *THRB* gene, encoding for thyroid hormone receptor (TR)β1 and β2, lead to autosomal dominant macular dystrophy (ADMD). We report the detailed clinical characterization of two first-degree relatives with ADMD, heterozygous for *THRB* c.283+1G>A, and an unrelated ADMD patient with a novel variant, c.283G>C. The genomic and molecular consequences of both variants were studied.

**Methods:** gDNA and mRNA were obtained from leukocytes. Clinical characterization included biochemistry, bone density and body composition, ECG, echocardiography, ultrasound, audiometry and color-vision. *In vitro* assays investigated TR function and DNA binding.

**Results:** The patients manifested no resistance to thyroid hormone beta (RTHβ) and had normal FT4 and TSH. Detailed studies in two patients showed no goiter, tachycardia, hypercholesterinemia or hepatic steatosis. Hearing was not impaired. Both had impaired color vision and reduced bone density. RT-PCR from all three patients revealed skipping of exon 4 exclusive to TRβ1, producing a deletion of 87 amino acids in the N-terminal domain (TRβ1^ΔNTD^). *In vitro*, DNA-binding affinity of TRβ1^ΔNTD^ to DR4-TRE with or without RXRα was comparable to TRβ1^WT^. Surprisingly, TRβ1^ΔNTD^ was transcriptionally twice more active than TRβ1^WT^ with a similar EC_50_ for T3, demonstrating gain-of-function of TRβ1^ΔNTD^. *THRA* expression in leukocytes was increased by 3-fold compared to unrelated controls and different from RTHβ patients.

**Conclusion:** These *THRB* splice site variants produce TRβ1 exon 4 skipping, resulting in a gain-of-function mutant, TRβ1^ΔNTD^. This explains the dominant ADMD phenotype devoid of RTHβ and suggests a TRβ1 gain-of-function syndrome.

## Introduction

Thyroid hormone (TH) is essential for organ development and metabolic homeostasis. The effects of 3,3′,5- triiodo-L-thyronine (T3), the active TH, are mediated by TH receptors (TRs) α and β. TRβ is expressed in two isoforms that differ in their N-termini, TRβ1 and TRβ2. TRβ1 is expressed in hepatocytes, but also in retina and anterior eye tissues, e.g. iris and ciliary body, whereas TRβ2 is restricted to the hypothalamus, pituitary, cochlea, and retina^1, 2^. TRβ2 regulates the hypothalamus-pituitary-thyroid (HPT) axis and, thus, the concentrations of TSH and TH (triiodothyronine, T3, and thyroxine, T4).

TRs consist of an N-terminal domain (NTD), a conserved DNA-binding domain (DBD), through the hinge region connected to the ligand-binding domain (LBD). The LBD and hinge regions are common for TRβ1 and 2 and their loss-of-function pathogenic variants are well characterized as the cause of autosomal dominant resistance to thyroid hormone beta (RTHβ)^3, 4^.

A novel *THRB* variant outside the LBD and hinge region (c.283+1G>A) was recently reported in association with autosomal dominant macular dystrophy (ADMD) as the major clinical feature in three Spanish families^5^. In 2025, two more Spanish families with the same c.283+1G>A variant and a second variant in another Spanish family, c.283G>A, affecting an adjacent nucleotide, were also reported^6^. We identified the c.283+1G>A variant in two first-degree relatives of a German family with macular degeneration and high intrafamilial variability of the retinal phenotype^7^.

The affected patients with ADMD were euthyroid and did not present the typical RTHβ phenotype despite the *THRB* pathogenic variants, which raised several questions: How do these variants lead to a phenotype? The *THRB* c.283+1G>A and c.283G>A variants alter the sequence of the donor splice site at the 3’ end of the N-terminal TRβ1 exclusive exon 4 and, presumably, affect *THRB* RNA splicing. What are the molecular consequences of the splice site mutation or, more precisely, what are the splicing products in the patients? How is TRβ1 function altered and how can the autosomal dominant phenotype expression be explained?

To address these questions, we characterized the clinical features of two patients harboring the *THRB* c.283+1G>A variant and a patient with a novel *THRB* c.283G>C variant as well as the effects on *THRB* splicing and TRβ function *in vitro*. Based on our results, we propose a new mechanism for these N-terminal TRβ1 pathogenic variants.

## Materials and Methods

### Ethical approval and informed consent statements

The Institutional Review Board of the University of Chicago approved this study (10-696-B), and the Ethics Committee of the University of Duisburg-Essen waived the requirement for ethics approval (25-12872-BO). All participants provided written informed consent prior to enrolment in the study.

### Clinical investigations

Two first-degree related adult patients from Germany with the *THRB* c.283+1G>A variant (Patients A and B), mother and son, were referred for further endocrinological evaluation. Clinical characterization included laboratory tests, bone mineral density and body composition (Dual-energy X-ray absorptiometry [DXA], SECA scale), thyroid ultrasound, liver ultrasound including shear wave and FibroScan, 12-lead ECG, echocardiography, pure-tone audiometry and color-vision testing (Ishihara plates and Farnsworth-15D). In addition, a blood sample, fundus imaging and optical coherence tomography (OCT) were obtained from a patient residing in New Zealand harboring a novel *THRB* c.283G>C variant (Patient C).

### Molecular genetics

gDNA extraction from whole blood of patients A, B, C and three unrelated normal controls was performed following kit manuals (NEB, Cat #T3010S; Zymo Research, Cat #D7003). Primers flanking exon 4 of *THRB* were used and the PCR product was analyzed by Sanger sequencing. For detection of splice variants white blood cells (WBCs) were separated from patient’s whole blood using BioColl^®^ (Bio&Cell, Nuremberg, Germany) and mRNA was extracted, and cDNA was synthesized as previously described^8^. All subjects’ cDNA flanking exon 3-7 was amplified by using ZymoTaq DNA Polymerase system and submitted to sequencing^8^.

For qPCR analysis, all gene expressions were normalized by geometric mean of four housekeeping genes, *POLR2A*, *ACTB*, *GAPDH* and *PPIA.* All primer sequences are shown in suppl. Table S1).

### Expression constructs and cell culture

Expression constructs for deletion of 87 amino acids, p.Glu8_Lys94del (TRβ1^ΔNTD^), p.Cys93_Gly95delinsTrp and p.G95R TRβ1 variants and IR0 (inverted repeat 0) TRE-luciferase reporter plasmid were generated by site-directed mutagenesis (Q5^®^ Site-Directed Mutagenesis Kit, NEB) using human TRβ1-WT in pSBbi-Neo or DR4-luciferase reporter in pcDNA3 as templates, respectively. For *in vitro* transcription, TRβ1 constructs were subcloned by Gibson Assembly (NEBuilder^®^ HiFi DNA Assembly Master Mix, NEB) into pcDNA3.1 expression vector downstream of the T7 promoter. RL-TK reporter plasmid (Renilla luciferase) and RXRα co-expression plasmid (pcDNA3.1-RXRα-C-HA) were used as indicated.

### Electrophoretic mobility shift assay (EMSA)

EMSA was performed as previously described with minor modifications^9^. Briefly, equal amounts (3.5 µl) of each *in vitro* translated TRβ1 variant and RXRα were preincubated in reaction buffer (20 mM HEPES, 50 mM KCl, 1 mM EDTA, 1 mM DTT, 1 mg/ml fatty acid-free BSA, 66 ng/µL poly-dI:dC and 50% glycerol) for 10 min at 20°C. The hybridized DR4- or IR0-probe was added to a final concentration of 10 nM and incubated for 30 min at 20°C in the dark. Samples were run on a 2.5% TBE-agarose gel at 200 V for 40 min and visualized afterwards using the VersaDoc imaging system (Bio-Rad, USA). For TH treatment, T3 (100 nM) was added during the 30 min incubation period.

### Luciferase reporter assay

Dual-luciferase reporter assay was performed as previously described using HEK293 cell with minor modifications^8^. Cells were co-transfected with plasmids (TRE-driven firefly luciferase reporter [DR4 or IR0], Renilla luciferase control, TRβ1^WT^ or TRβ1^ΔNTD^ ± RXRα) using PEI (Polyethyleneimine). 24h post-transfection cells were treated with solvent or T3 (0–100 nM). After 24h luciferase signals were measured on a GloMax Discovery (Promega, Madison, WI, USA).

### Statistical and computational analysis

If not otherwise indicated, data are shown as mean ± SEM. Statistical analysis was done with GraphPad Prism 10 and p<0.05 was considered significant. SpliceAI was used via https://spliceailookup.broadinstitute.org reporting a Δ score between 0 to 1 as the probability of affecting splicing^10^. TRβ1^WT^ and TRβ1^ΔNTD^ protein structure were modelled with AlphaFold 3^11^.

## Results

### Clinical Phenotype

#### Patients A and B

The proposita (Patient A), a woman in her late 60’s had a chief complaint of impaired vision for many years. She had corrective since her early 30’s. After the diagnosis of macular degeneration was made, whole-genome sequencing to determine the cause for ADMD revealed the c.283+1G>A variant in the *THRB* gene^7^. Her past medical history includes cataract surgery with lens replacement in both eyes (early 60’s) and breast cancer with surgery and tamoxifen treatment (late 40’s). At that time osteoporosis was diagnosed and treated with bisphosphonate for 10 years, followed by denosumab until present together with Vit. D 20.000 IU/week. After detection of the variant, her son (Patient B) was studied and the same *THRB* variant was found. He was diagnosed with vitelliform macular degeneration in his early 40’s. He had corrective lenses for about 20 years and noticed a deterioration of vision in the last 5 years. He reported no relevant past medical history and no current medications.

They were of normal height and weight (Table 1). They were euthyroid (Table 1) and the ratio of FT4 to FT3 was not abnormal. Thyroid autoantibodies (anti-TG, TPO, TSHR) were negative. Ultrasound showed small or normal thyroid glands (4.5 ml [mother], 9 ml [son]), of normal echogenicity and without nodules. Total cholesterol as well as HDL- and LDL cholesterol and triglycerides were within the reference range (Table 1). Abdominal ultrasound showed no severe abnormalities, especially no signs of hepatic steatosis. Non-invasive techniques further supported the absence of hepatic steatosis. Controlled attenuation parameter (CAP) was below the threshold for hepatic steatosis (<275 dB/m) and stiffness was not increased (<8 kPa) (Table 1). Bone mineral density showed osteopenia in the mother and mild osteopenia in the son (Table 1). In a DXA body composition analysis, the mother’s whole body fat content was near the age-and sex-specific 50^th^ percentile (36.2%, Z score: 0.2). Her son’s fat content was higher for age and sex (30.0%, Z score: 1.8). Both patients had low or low normal skeletal muscle mass (Seca scale analysis; mother: 13.5 kg [15.4-22.0], son: 27.2 kg [25.5-33.6]).

**Table 1:** Clinical characteristics of the two THRB c.283+1G>A patients.

|  | Patient A | Patient B |  | Reference range |
| --- | --- | --- | --- | --- |
| height | 160 | 175 | cm |  |
| weight | 53 | 75 | kg |  |
| BMI | 20,7 | 24,5 | kg/m<sup>2</sup> |  |
| TSH | 2,15 | 0,86 | mU/l | 0,55-4,78 |
| FT4 | 17,0 | 15,2 | pmol/l | 11,5-22,7 |
| TT4 | 95,0 | 88,0 | nmol/l | 58-141 |
| FT3 | 6,2 | 5,8 | pmol/l | 3,5-6,5 |
| TT3 | 1,70 | 1,74 | nmol/l | 0,92-2,79 |
| rT3 | 198,1 | 178,4 | pg/ml | 90-215 |
| total cholesterol | 194 | 168 | mg/dl |  |
| HDL | 78 | 49 | mg/dl | >40 |
| LDL | 101 | 114 | mg/dl |  |
| triglycerides | 57 | 76 | mg/dl |  |
| Liver: CAP | 217 | 174 | dB/m | <275 |
| Liver: stiffness | 5,3 | 4,0 | kPa | <8 |
| EF (echocardiography) | 65% | 55% |  |  |
| T-Score L1-L4 (DXA) | -1,3 | -1,3 |  |  |
| T-Score total hip (DXA) | -2,0 | 0,0 |  |  |

Heart rate was not increased with 63 bpm (mother) and 59 bpm (son). Cardiac function was not impaired (Table 1). Dimensions of the cardiac walls were normal, no pericardial effusion. Audiometry showed normal hearing for the respective age for both ears in both patients (Fig. 1A). Otoacoustic emissions were present without abnormalities across all frequencies. We studied color vision and both patients showed abnormal results in the Ishihara test. The proposita recognized none of 12 plates with either eye. Her son recognized 2/12 plates with the right eye and none with the left eye. The Farnsworth-15D test revealed mild deviations (indices within 2 standard deviations but trending high; Table 1) (Fig. 1B).

**Figure 1:**
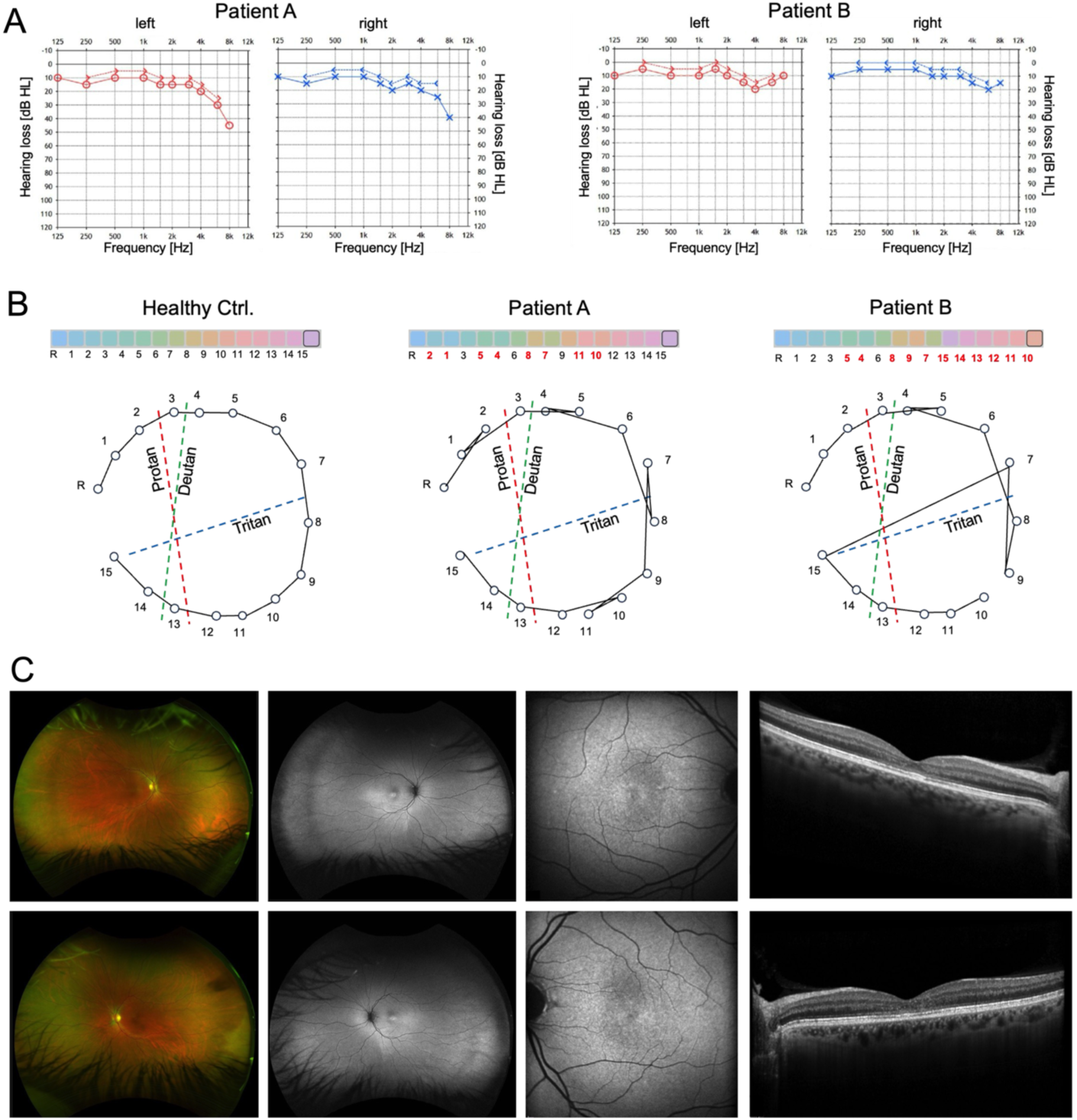
Audiometry, color vision and retinal imaging. **A** Patients A and B: Bilateral audiometry was performed in a range from 125 Hz to 8 kHz. The audiogram was normal and according to age for both patients. **B** Patients A and B: Farnsworth desaturated 15 color vision test revealed mild color vision defects. **C** Patient C: Wide-field false-color fundus photography (first column), 55 degree blue-light fundus autofluorescence (second column), 30 degree blue-light fundus autofluorescence imaging (FAF) (third column) and horizontal spectral-domain optical coherence tomography line scan (SD-OCT) (last column).

#### Patient C

An unrelated female patient in her early 50’s (Patient C) from a three-generation with ADMD was found to have ADMD with a sawtooth appearance of the ellipsoid line and outer photoreceptor segments in OCT (Fig. 1C). In exams three and five years later, photoreceptor layers showed progressive thinning. During this time, the patient experienced increasing difficulty with reading (right visual acuity 6/7.5-2 unaided, left visual acuity 6/9-2, both eyes 6/7.5-1). She was euthyroid (TSH 1.2 U/l [0.27-4.20]). She has a history of constipation with several episodes of impaction and has required iron infusions for iron deficient anemia due to menorrhagia. She had fibroadenomas in both breasts that were removed when she was a young adult. In her early 50’s she was found to have PT1cN0 grade II invasive ductal carcinoma (ER/PR positive, HER2 negative breast cancer). She was treated with radiotherapy and tamoxifen for 5 years. In 2025, the breast cancer recurred, and she is actively undergoing treatments. The patient’s mother, now in her early 80’s, was diagnosed with eye disease in her early 40’s and had an ovarian mucinous borderline tumor in her early 50’s. She was euthyroid on levothyroxine for autoimmune hypothyroidism (TSH 2.5 U/l [0.3-4.0]). The patient’s teenage son was found to have macular pigmentary changes as a school-age child. The affected persons of this family did not report any hearing issues.

### Both *THRB* c.283+1G>A and c.283G>C donor splice site variants lead to skipping of N-terminal TRβ1-specific exon 4

Sequencing of gDNA confirmed the c.283+1G>A donor splice site pathogenic variant in the *THRB* expressing TRβ1 of patients A and B and a novel c.283G>C in the last nucleotide of TRβ1 exon 4 in Patient C (Fig. 2A).

**Figure 2:**
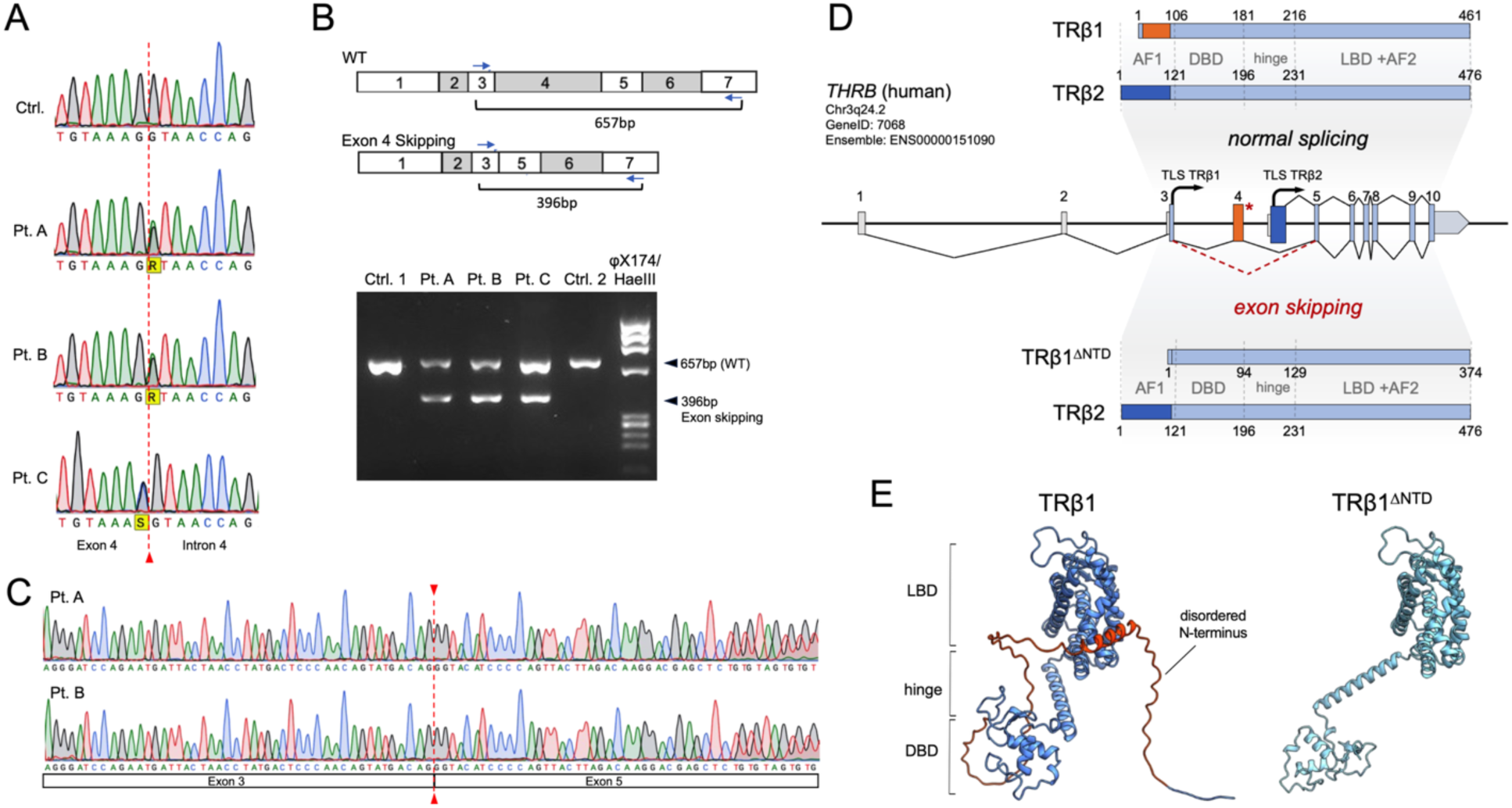
Consequences of the *THRB* exon 4 splice site mutation. **A** Sequencing of THRB from gDNA (whole blood): healthy wild-type control (Ctrl.), heterozygous c.283+1G>A mutation in two patient (Pt. A and B) and heterozygous c.283G>C mutation in patient C. **B** THRB transcript analysis: PCRs on cDNA reverse transcribed from leukocyte RNA from Pt. A and B. **C** Sequencing of the PCR products from B (lower bands). **D** Genomic structure of WT and mutant human *THRB* and consequences for TRβ1 and TRβ2 proteins (exon numbering for hTRβ1^12^). **E** Modeling of TRβ1 (UniProt: P10828) and TRβ1^ΔNTD^ with AlphaFold 3^11^ shows that skipping of exon 4 results in an almost complete loss of the intrinsically disordered N-terminal domain (red) of TRβ1. LBD = ligand binding domain, hinge = hinge region, DBD = DNA binding domain.

Computational splice site analysis predicted a high probability for loss of this donor splice site due to both mutations (Δ scores for donor loss of 0.98 and 0.80, respectively)^10^. To determine the consequences of these variants in material from the patients, we amplified cDNA generated from WBC mRNA, which yielded two products, one of the expected length for the WT TRβ1 allele and a shorter band (Fig. 2B). The latter lacked 261 nucleotides, corresponding to the 87 amino acids encoded by exon 4 of TRβ1, glutamic acid 8 through lysine 94 (E8-K94) (Fig. 2B,C). Thus, in patients’ samples, both variants resulted in a heterozygous in-frame deletion of 87 amino-acids from the NTD through skipping of exon 4 of TRβ1 (Fig 2D), a loss of 82% of the NTD (87/106 amino acids, TRβ1^ΔNTD^). The deletion of exon 4 removes most of the TRβ1 intrinsically disordered region (IDR) (Fig. 2E). The phase 1 transition to exon 5 is retained, which changes the amino acid Glu (GAA) to Gly (GGG) at the new exon 3 to 5 junction and preserving the amino acid, Gly, at the WT 4 to 5 junction. The percentage of the heterozygous N-terminal deletion variant (TRβ1^ΔNTD^), common to both pathogenic variants, in the total amount of TRβ1 was 63% (Patient A), 57% (Patient B) and 48% (Patient C), demonstrating that exon skipping is the major result and near complete in mRNA from the mutant alleles.

In addition to skipping of exon 4, results from an *in vitro* minigene splice assay^6^ had suggested deletion of two amino acids and an insertion of one amino acid (p.Cys93_Gly95delinsTrp) as a consequence of the c.283+1G>A and c.283G>A donor splice site variants. For the c.283G>A variant, an additional a missense variant (p.G95R) was predicted. In mRNA from the patients, we did not find transcripts with the putative missense variant. When selectively amplifying transcripts containing exon 4 using a forward primer in exon 4 and a reverse primer in exon 7, minimal amounts of transcripts encoding for the p.Cys93_Gly95delinsTrp were detected only in patients A and C (on average <6%) and not in patient B. *In vitro* transcribed TRβ1 variants p.Cys93_Gly95delinsTrp and p.G95R demonstrated normal transactivation in luciferase reporter assays and TRE binding in EMSA (Suppl. Fig. 1).

### Skipping of exon 4 does not interfere with TRβ1 DR4 TRE binding or homo- and heterodimerization

Because of the structural proximity of the N-terminus to the DBD we performed EMSA to investigate whether deletion of exon 4 affects DNA binding or formation of homo- and heterodimers. In the absence of the main TR binding partner RXRα, the full-length WT TRβ1 binds to DR4 and IR0 as a homodimer (Fig. 3A). The TRβ1^ΔNTD^ mutant also bound as a homodimer to DR4. However, TRβ1^ΔNTD^ did not bind to IR0 (Fig. 3A). With RXRα present, both TRβ1^WT^ and TRβ1^ΔNTD^ formed heterodimers on DR4 and IR0 (Fig. 3A).

**Figure 3:**
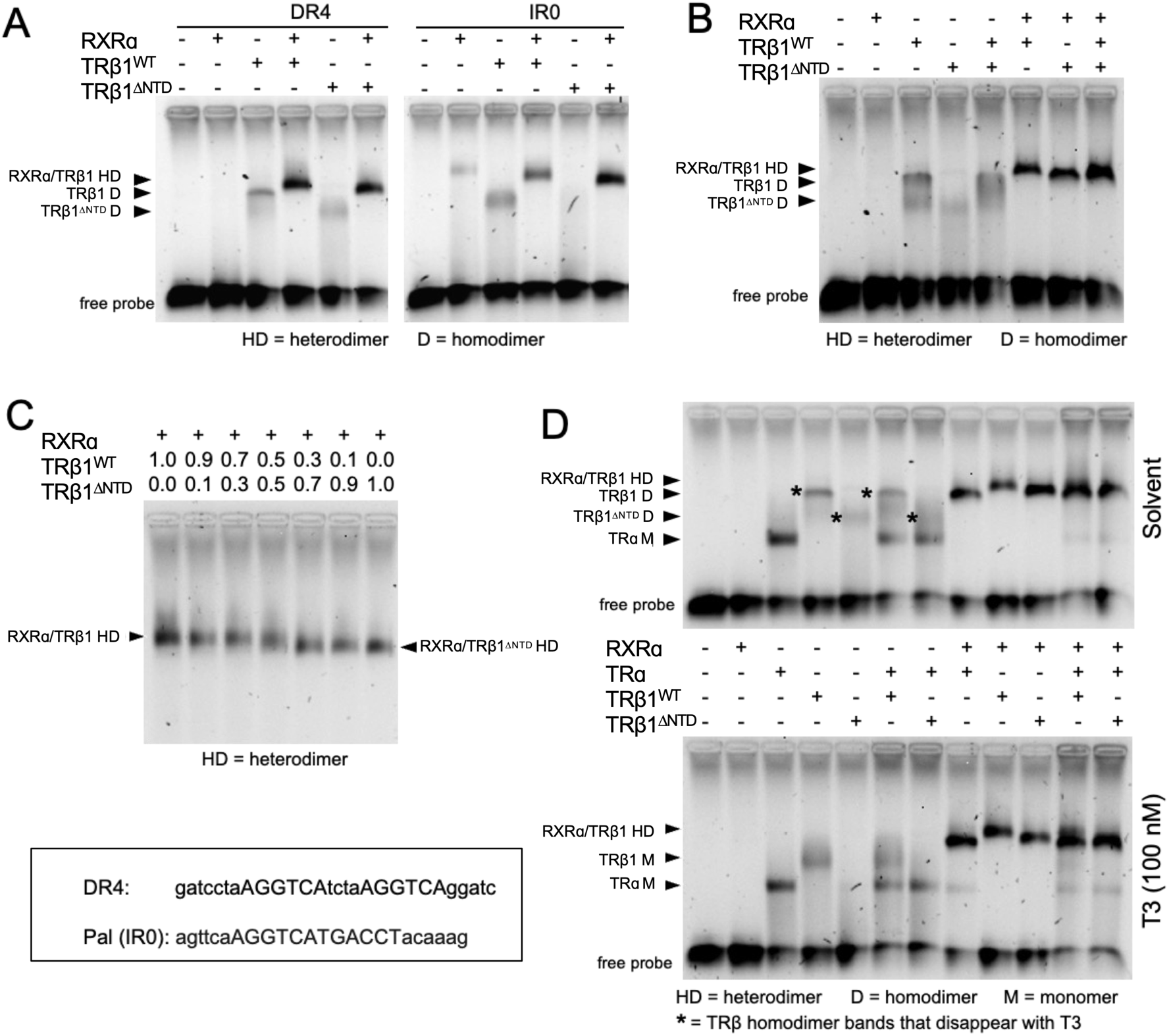
Electrophoretic mobility shift assay (EMSA). **A** TRβ1^WT^ and TRβ1^ΔNTD^ with or without RXRα, were incubated with fluorescent EMSA probes (Cy5) of the most prominent TREs, DR4 and IR0 (box). **B** To test for TRβ1^WT^/TRβ1^ΔNTD^ heterodimers, equal amounts of TRβ1^WT^ and TRβ1^ΔNTD^ were incubated with DR4-Cy5 in presence or absence of RXRα. **C** Increasing amounts of TRβ1^ΔNTD^ were to test for interference with TRβ1^WT^/RXRα heterodimer formation. **D** To study a possible effect of T3 on dimerization and DNA binding we performed without (solvent) and with T3 (100 nM) and included TRα.

As the patients are heterozygous for WT and mutant TRβ1, we next used equal amounts of TRβ1^WT^ and TRβ1^ΔNTD^ without and with RXRα on a DR4. TRβ1^WT^ and TRβ1^ΔNTD^ seemed not to form a WT/mutant dimer, but together with RXRα and using alternating amounts we observed a broad band, suggestive of a composite of TRβ1^WT^/RXRα and TRβ1^ΔNTD^/RXRα heterodimers (Fig. 3B & C).

We investigated whether T3 influences TRβ1^ΔNTD^ DNA complex formation and whether TRα DNA binding is affected by TRβ1^ΔNTD^. For both TRβ1^WT^ and TRβ1^ΔNTD^, homodimers disappeared with T3 treatment, but TRβ1^ΔNTD^ did not form monomers like the WT (Fig. 3D).

In absence of RXRα and T3, TRα bound to DR4 as a monomer. With or without RXRα and ±T3, the DNA-binding pattern of TRα and TRβ1^ΔNTD^ together was the same as alone, suggesting no interference with each other’s DNA binding ability (Fig. 3D).

### TRβ1^ΔNTD^ manifests gain-of-function *in vitro* using a TRE luciferase reporter

The transcriptional activity of TRβ1^ΔNTD^ was studied in luciferase assays. T3 treatment of TRβ1^ΔNTD^ led to an almost two-fold increase in luciferase activity compared to TRβ1^WT^ on the DR4 reporter, co-expressed with RXRα (p<0.01) (Fig. 4A), indicating that loss of exon 4 does not lead to loss-of-function, but rather gain-of-function. T3 EC_50_ was similar for TRβ1^WT^ and TRβ1^ΔNTD^ (WT 16.2+0.1 nM; ΔNTD 14.1±0.1 nM) (Fig. 4B). When normalizing to the plateau reached with T3 (10^-6^ M), the dose response curves for TRβ1^WT^ and TRβ1^ΔNTD^ were superimposable (Fig. 4B, lower panel). The increased transcriptional activity of TRβ1^ΔNTD^ did not depend on RXRα (Fig. 4C).

**Figure 4:**
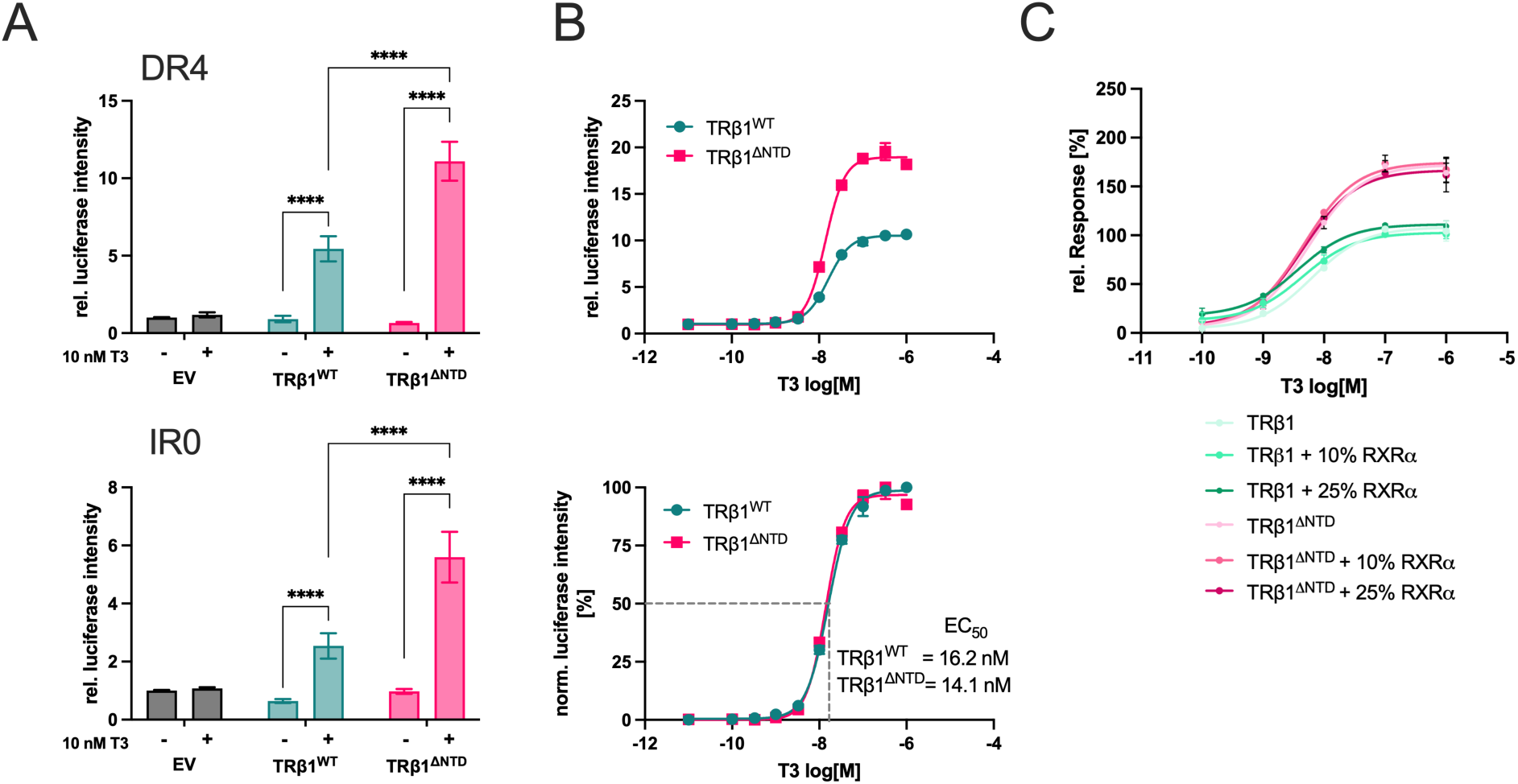
Luciferase reporter assay for TRβ1^WT^ and TRβ1^ΔNTD^. **A** Dual-luciferase assay with DR4- and IR0-reporter, stimulated with 10 nM T3 for 24h (n=3; One-way ANOVA with Tukey post-hoc test; ****p<0.0001). **B** T3 dose-response curves for TRβ1^WT^ and TRβ1^ΔNTD^ and estimation of EC_50_ values. **C** T3 dose-response of co-expression with RXRα from 0% to 25% of TRβ1 (WT and ΔNTD).

### Increased expression of *THRA* in blood samples of patients with *THRB* c.283+1G>A and c.283G>C

As the clinical evaluation seemed to find some features involving TRɑ predominant tissues, we assessed the level of expression of *THRA* isoforms in peripheral blood leucocytes from these three patients compared to unrelated normal controls. We also assessed *THRA* expression in 10 affected members vs 11 unaffected first-degree relatives from five families with RTHβ, caused by previously reported missense pathogenic variants p.A317T in two of the families and p.P452S, E460K and A279T each in one family. Total *THRA* expression was 3-fold higher in samples from individuals heterozygous for *THRB* c.283+1G>A and c.283G>C compared to expression in five unrelated controls, with a mild, not significant increase in the expression of *THRA1* isoform of 1.6-fold, and a significant 1.8-fold increased expression of the *THRA2* isoform (Fig. 5A). In contrast, in patients with RTHβ patients compared to unaffected relatives, *THRA1* and *THRA2* expression was decreased to 53% and 42% (Fig. 5B).

**Figure 5:**
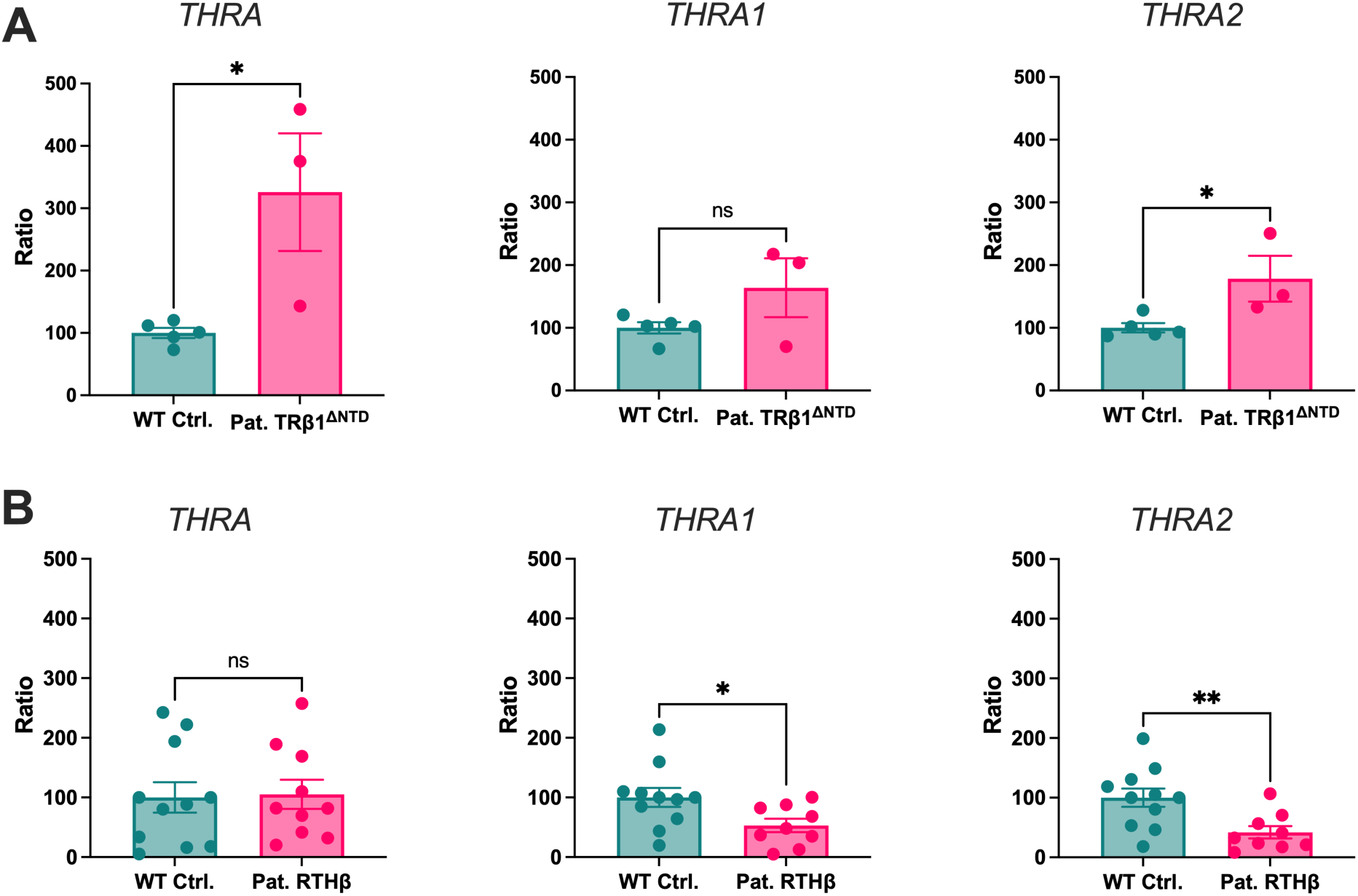
Comparison of *THRA* isoform expression in patients with TRβ1^ΔNTD^ and RTHβ. Expression of total *THRA*, *THRA1* and *THRA2* in **A** healthy WT individuals and patients with TRβ1^ΔNTD^ mutations, and **B** patients with RTHβ and their first-degree relatives was analyzed by qPCR of cDNA from whole blood. (A: n=3-5; Unpaired t-test; ns= not significant, *p<0.05; B: n=11; Unpaired t-test; ns= not significant, *p<0.05, **p<0.01)

## Discussion

Our data show that the *THRB* c.283+1G>A and the novel c.283G>C splice site variants in patients with ADMD affect exclusively TRβ1 and lead to deletion of 87 amino acids due to exon 4 skipping, resulting in an N-terminal deletion variant, TRβ1^ΔNTD^. Importantly, transcriptional activity of TRβ1^ΔNTD^ is increased about two-fold compared to WT TRβ1. This TRβ1 gain-of-function explains why patients with these variants present with a dominant phenotype that is different from the loss-of-function phenotype produced by mutations in the LBD of the TRβ.

Patients with ADMD have been recently described to harbor the N-terminal *THRB1* specific c.283+1G>A and the adjacent c.283G>A pathogenic variants^5, 6^. We report a new family with two affected individuals (Patient A and B) harboring the *THRB* c.283+1G>A heterozygous pathogenic variant, and another unrelated individual (Patient C) with a novel variant in the last coding nucleotide of the TRβ1-specific exon 4, c.283G>C, a different nucleotide change than previously reported in this position^6^. Thus, three heterozygous N-terminal pathogenic *THRB* variants in the donor splice site of TRβ1-specific exon 4 have been identified in nine different families with ADMD^5-7^.

Analysis of cDNA generated from blood samples of the three patients’ reported herein, revealed that skipping exon 4 was the main consequence for both donor splice site variants, *THRB* c.283+1G>A and the novel variant c.283G>C, with minimal, if any, transcripts encoding for p.Cys93_Gly95delinsTrp, and no transcripts encoding the missense p.Gly95Arg. Exon 4 is located upstream from the TRβ2-specific promoter and translation start site and is therefore TRβ1 exclusive^13^. Skipping exon 4 results in deletion of 87 amino acids, E8-K94, encoding for >80% of the TRβ1 N-terminal IDR (TRβ1^ΔNTD^). TRβ1^ΔNTD^ was able to bind a DR4 TRE as homodimer and heterodimer with RXRɑ. Interestingly, the deletion of TRβ1 exon 4 did not lead to impaired TRβ1 action and resistance to TH. In contrast, the truncated TRβ1^ΔNTD^ was transcriptionally about twofold more active than TRβ1^WT^. Deletion of exon 4 therefore led to TRβ1 gain-of-function as a new pathogenic principle for TRβ with clinical consequences.

Retinopathy with macular dystrophy appears to be the clinical hallmark of the TRβ1-specific gain-of-function variant. How could TRβ1 gain-of-function explain this phenotype? The retina is a T3-dependent tissue, and T3 determines cone photoreceptor precursor development into M- and S-cones^14^. While TH is necessary for proper photoreceptor development, too much TH action due to hyperthyroidism is detrimental and leads to loss of cones and rods. In mice with a genetic retinopathy background and even in WT mice, impaired retinal function, caused photoreceptor degeneration, and induced oxidative stress, cell death, and induced genes involved in cellular stress, necroptosis, and inflammation^15, 16^. These results from mice support a role for increased TRβ signaling in the pathogenesis of retinal degeneration. Epidemiological studies suggest that the detrimental effect of excess TH action in mice can be translated to humans. Higher FT4 concentrations and overt hyperthyroidism were associated with increased risk of age-related macular degeneration^17, 18^. As TRβ1 is the TRβ isoform expressed after fetal development^2^ it is reasonable to assume that constantly increased TH action due to TRβ1 gain-of-function in TRβ1^ΔNTD^ promotes retinal degradation similar to high TH concentrations.

Color vision has not previously been reported or specifically studied in patients with the *THRB* c.283+1G>A or c.283G>A variants^5, 6^. However, color vision was impaired in patients A and B. It is possible that these defects are due to cone loss as part of the retinopathy. As cone development is regulated by TH and the TRs, increased TH action may also disturb cone development or differentiation. Whether cone loss or disturbed cone development, or both, contribute to the color vision impairment found in the present patients remains to be determined.

The phenotype of ADMD with *THRB* variants and RTHβ is very different. In RTHβ, pathogenic variants in the ligand-binding and hinge domains are shared by both TRβ1 and TRβ2 and manifest with high serum total and free TH with normal or high TSH concentration, the characteristic biochemical features of RTHβ. This is explained by reduced sensitivity to TH in TRβ-expressing tissues (impaired feedback in the HPT axis, reduced TH action in the liver), while TRα-expressing tissues, especially heart, gut and bone, retain normal sensitivity and respond to the elevated TH serum concentrations. RTHβ patients present with goiter, impairment of color vision, tachycardia, dyslipidemia, and hepatic steatosis^3, 4, 19-21^. In contrast, information from the initial reports of ADMD patients with *THRB* variants indicated that a serum thyroid phenotype is lacking^5-7^. The phenotype of the patients reported herein with the *THRB* c.283+1 G>A and c.283G>C variants is different from that of patients with RTHβ. Most obviously, they are euthyroid as the pituitary TRβ2 is not affected. Furthermore, our two patients harboring *THRB* c.283+1G>A gain-of-function mutation showed no tachycardia and did not present with hepatic steatosis or elevated serum lipids. In RTHβ, the dyslipidemia and development of hepatic steatosis result from resistance to TH action and reduced lipolysis and beta oxidation in hepatocytes. The TRβ1^ΔNTD^ mutant does not confer resistance to TH, which explains the absence of dyslipidemia and hepatic steatosis in the present patients. The difference in TRβ function, TRβ1 gain-of-function due to TRβ1 exon 4 deletion in TRβ1^ΔNTD^ vs. TRβ1 and TRβ2 loss-of-function in RTHβ, provides an explanation for the phenotype difference. Importantly, TRβ1 gain-of-function also explains the dominant inheritance observed in ADMD families with these heterozygous *THRB* pathological variants.

Although TH and TRβ1 are relevant for auditory development^22^, hearing was not overtly affected in the present patients. Bone mineral density was reduced in the trabecular bones of the lumbar spine (L1-L4), a notable finding in the male patient B without obvious risk factors for osteopenia. Although expression of TRɑ1 is at least 10-fold greater than TRβ1 in bone, TRβ1 also mediates TH effects^23^. Specifically, TH induced trabecular bone loss in TRɑ-deficient mice^24^. Constantly increased TRβ1 signaling could, therefore, induce osteopenia, but more patients’ data are needed to determine whether osteopenia is a clinical feature of TRβ1 gain-of-function. A potential cross talk with TRɑ was also considered. Mean expression of total *THRA* was 3-fold higher in peripheral blood leucocytes from the three patients investigated here compared to unrelated controls with significant increase of *THRA2* expression. This was in contrast with the overall normal *THRA* expression in blood samples from RTHβ patients compared to unaffected relatives, and significant decrease in *THRA2* expression. This result further supports that the mechanism and syndrome reported here and RTHβ are very different, almost opposite.

How can loss of a large part of the TRβ1 NTD lead to gain-of-function? The TRβ1 N-terminus could contain corepressor binding sites that are removed when exon 4 is deleted, which may reduce repression and increase transcription. The TRβ1 N-terminal region is an IDR. Negatively charged IDRs can mimic nucleic acids and compete with DNA for their own protein’s DBD, resulting in autoinhibition^25^, thought to reduce DNA-binding affinity but increase specificity. In the TRβ1^ΔNTD^ mutant, increased transcriptional activity could be explained by the large IDR deletion which may relieve autoinhibition. However, our competitive EMSA did not indicate altered DNA-binding of TRβ1^ΔNTD^. Therefore, the precise mechanism how the N-terminal deletion mediates TRβ1 gain-of-function has yet to be determined.

In summary, the present clinical and molecular data indicate that heterozygous TRβ1-specific exon 4 splice site variants in *THRB* cause a dominantly inherited phenotype through TRβ1 gain-of-function due to exon 4 skipping and deletion of almost the entire TRβ1 NTD. This mechanism differs from RTHβ loss-of-function pathogenic variants in the LBD or hinge region. Clinically, affected patients present with an undisturbed HPT axis and euthyroidism, but develop ADMD and impaired color vision likely reflecting chronic dysregulation of TRβ1 targets in the retina.

## Acknowledgements

We thank Andrea Jaeger for dedicated technical assistance.

## Authors’ Contributions

G.S.H., L.C.M.: Conceptualization, investigation, methodology, formal analysis, resources, visualization, funding acquisition, writing – original draft preparation, writing – review & editing. X.-H.L.: Investigations, methodology, formal analysis, visualization, writing – review & editing. E.A.M., P.H. & A.E.: Investigation, resources, writing – review & editing. D.F.: Investigation, funding acquisition, writing – review & editing. R.E.W., J.M.C. & J.C.: Methodology, visualization, writing – review & editing. A.L.V.: Investigation, resources, formal analysis, writing – review & editing. A.M.D. & S.R.: Conceptualization, investigation, methodology, formal analysis, resources, funding acquisition, writing – review & editing.

## Statements and declarations

### Ethical considerations

The Institutional Review Board of the University of Chicago approved this study (10-696-B) and the Ethics Committee of the University of Duisburg-Essen waived the requirement for ethics approval (25-12872-BO).

### Consent to participate

All participants provided written informed consent for participation in the study.

### Consent for publication

All participants provided written informed consent for publication of their data.

### Data availability

All data generated or analyzed during this study are included in this article and its supplementary information files.

## Tables and Figures

**Supplementary Table 1:**
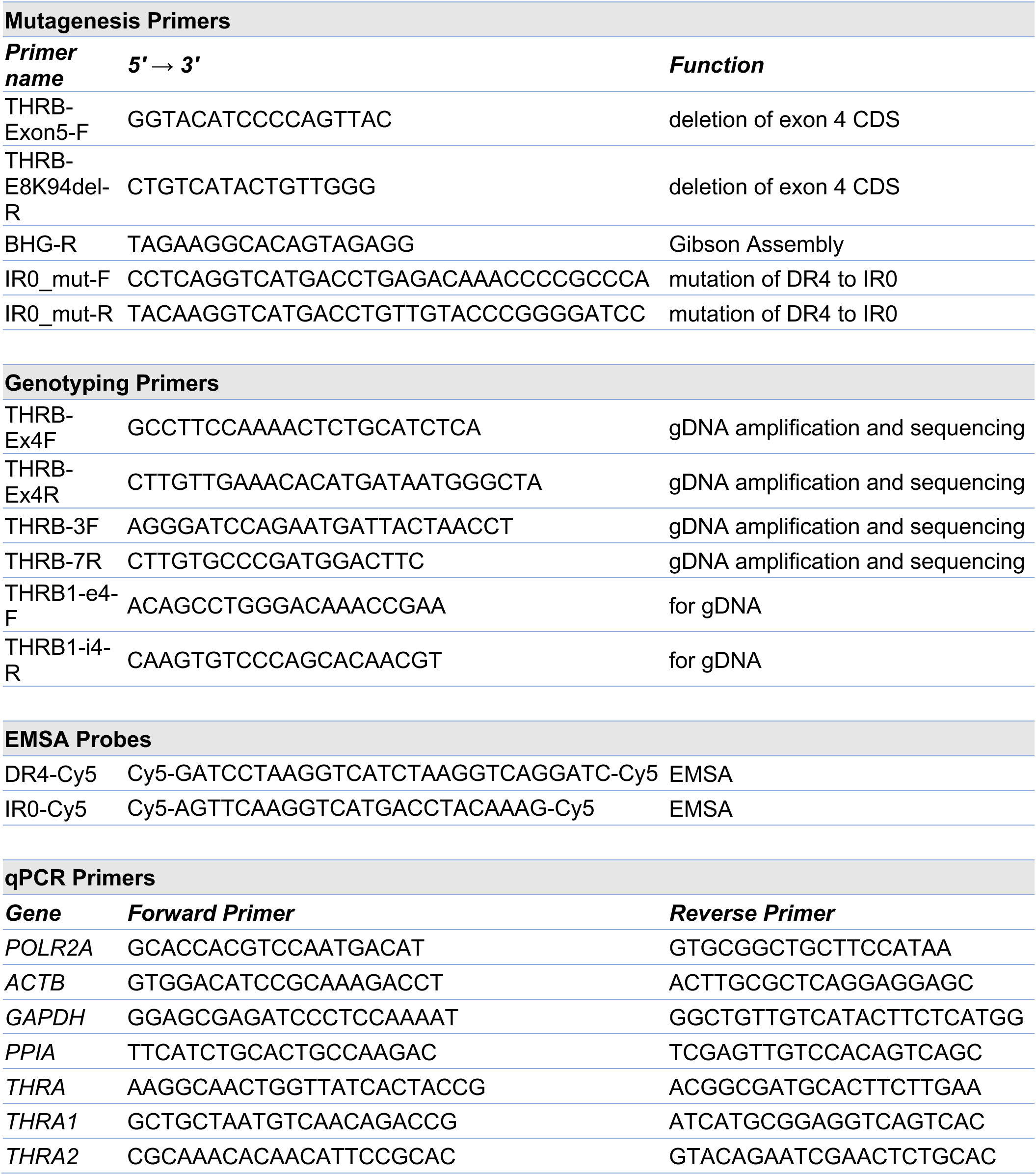
List of primers and EMSA probes used in this study.

**Figure S1:**
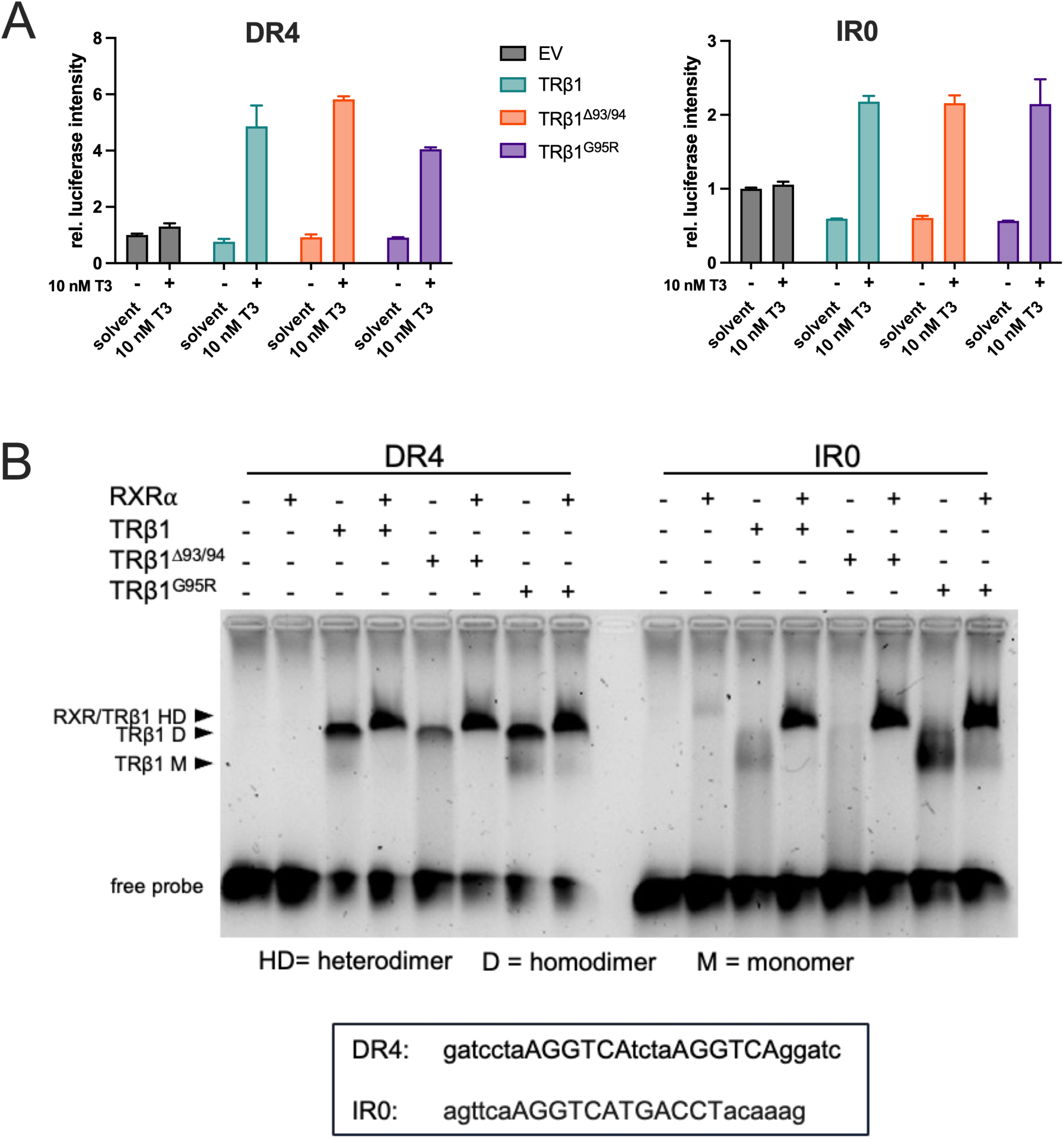
Luciferase assay and EMSA for TRβ1 mutants G95R and Δ93/94. **A** Dual-luciferase assay with DR4- and IR0-reporter, stimulated with 10 nM T3 for 24h. **B** TRβ1^WT^, TRβ1^Δ93/94^ (p.Cys93_Gly95delinsTrp) and TRβ1^G95R^ with or without RXRα, were incubated with fluorescent EMSA probes (Cy5) of the most prominent TREs, DR4 and IR0 (box).

## References

1. Minakhina S, Bansal S, Zhang A, et al. A Direct Comparison of Thyroid Hormone Receptor Protein Levels in Mice Provides Unexpected Insights into Thyroid Hormone Action. Thyroid 2020 2020/03/04. DOI: 10.1089/thy.2019.0763.

2. Ng L, Liu H, Liu Y, et al. Biphasic expression of thyroid hormone receptor TRβ1 in mammalian retina and anterior ocular tissues. Frontiers in endocrinology 2023; 14: 1174600. 2023/04/11. DOI: 10.3389/fendo.2023.1174600.

3. Pappa T and Refetoff S. Resistance to Thyroid Hormone Beta: A Focused Review. Frontiers in endocrinology 2021; 12. Mini Review. DOI: 10.3389/fendo.2021.656551.

4. Dumitrescu AM and Refetoff S. The syndromes of reduced sensitivity to thyroid hormone. Biochimica et biophysica acta 2013; 1830: 3987–4003. 2012/09/19. DOI: 10.1016/j.bbagen.2012.08.005.

5. Fernandez-Suarez E, Gonzalez-Del Pozo M, Garcia-Nunez A, et al. Expanding the phenotype of THRB: a range of macular dystrophies as the major clinical manifestations in patients with a dominant splicing variant. Front Cell Dev Biol 2023; 11: 1197744. 20230721. DOI: 10.3389/fcell.2023.1197744.

6. Fernandez-Caballero L, Blanco-Kelly F, Swafiri ST, et al. Identification of new families and variants in autosomal dominant macular dystrophy associated with THRB. Scientific reports 2025; 15: 14904. 20250428. DOI: 10.1038/s41598-025-97768-9.

7. Mahler EA, Moeller LC, Wall K, et al. Mutation of the Thyroid Hormone Receptor Beta Gene (THRB) Causes Vitelliform Macular Dystrophy with High Intrafamilial Variability. Genes (Basel) 2025; 16: 1240. 20251020. DOI: 10.3390/genes16101240.

8. Hones GS, Rakov H, Logan J, et al. Noncanonical thyroid hormone signaling mediates cardiometabolic effects in vivo. Proceedings of the National Academy of Sciences of the United States of America 2017; 114: E11323–E11332. 2017/12/13. DOI: 10.1073/pnas.1706801115.

9. Roth L, Johann K, Hönes GS, et al. Thyroid hormones regulate Zfp423 expression in regionally distinct adipose depots through direct and cell-autonomous action. Cell reports 2023; 42: 112088. 20230207. DOI: 10.1016/j.celrep.2023.112088.

10. Jaganathan K, Kyriazopoulou Panagiotopoulou S, McRae JF, et al. Predicting Splicing from Primary Sequence with Deep Learning. Cell 2019; 176: 535–548 e524. 20190117. DOI: 10.1016/j.cell.2018.12.015.

11. Abramson J, Adler J, Dunger J, et al. Accurate structure prediction of biomolecular interactions with AlphaFold 3. Nature 2024; 630: 493–500. 20240508. DOI: 10.1038/s41586-024-07487-w.

12. Beck-Peccoz P, Chatterjee VK, Chin WW, et al. Nomenclature of thyroid hormone receptor beta-gene mutations in resistance to thyroid hormone: consensus statement from the first workshop on thyroid hormone resistance, July 10–11, 1993, Cambridge, United Kingdom. J Clin Endocrinol Metab 1994; 78: 990–993. DOI: 10.1210/jcem.78.4.8157732.

13. Jones I, Srinivas M, Ng L, et al. The thyroid hormone receptor beta gene: structure and functions in the brain and sensory systems. Thyroid 2003; 13: 1057–1068. Research Support, Non-U.S. Gov’t Research Support, U.S. Gov’t, P.H.S. Review 2003/12/04. DOI: 10.1089/105072503770867228.

14. Ng L, Liu Y, Cho YW, et al. Differentiation versus dysfunction: thyroid hormone, deiodinases and retinal photoreceptors. European thyroid journal 2025; 14 20250312. DOI: 10.1530/etj-24-0315.

15. Ma H, Thapa A, Morris L, et al. Suppressing thyroid hormone signaling preserves cone photoreceptors in mouse models of retinal degeneration. Proceedings of the National Academy of Sciences of the United States of America 2014; 111: 3602–3607. 20140218. DOI: 10.1073/pnas.1317041111.

16. Ma H, Yang F, York LR, et al. Excessive Thyroid Hormone Signaling Induces Photoreceptor Degeneration in Mice. eneuro 2023; 10: ENEURO.0058–0023.2023. DOI: 10.1523/eneuro.0058-23.2023.

17. Chaker L, Buitendijk GH, Dehghan A, et al. Thyroid function and age-related macular degeneration: a prospective population-based cohort study--the Rotterdam Study. Bmc Med 2015; 13: 94. 20150423. DOI: 10.1186/s12916-015-0329-0.

18. Gopinath B, Liew G, Kifley A, et al. Thyroid Dysfunction and Ten-Year Incidence of Age-Related Macular Degeneration. Investigative ophthalmology & visual science 2016; 57: 5273–5277. DOI: 10.1167/iovs.16-19735.

19. Campi I, Cammarata G, Bianchi Marzoli S, et al. Retinal Photoreceptor Functions Are Compromised in Patients With Resistance to Thyroid Hormone Syndrome (RTHβ). J Clin Endocrinol Metab 2017; 102: 2620–2627. DOI: 10.1210/jc.2016-3671.

20. Chaves C, Bruinstroop E, Refetoff S, et al. Increased Hepatic Fat Content in Patients with Resistance to Thyroid Hormone Beta. Thyroid 2021; 31: 1127–1134. 2020/12/24. DOI: 10.1089/thy.2020.0651.

21. Moran C, McEniery CM, Schoenmakers N, et al. Dyslipidemia, Insulin Resistance, Ectopic Lipid Accumulation, and Vascular Function in Resistance to Thyroid Hormone β. J Clin Endocrinol Metab 2021; 106: e2005–e2014. 2021/02/02. DOI: 10.1210/clinem/dgab002.

22. Ng L, Kelley MW and Forrest D. Making sense with thyroid hormone--the role of T(3) in auditory development. Nature reviews Endocrinology 2013; 9: 296–307. 20130326. DOI: 10.1038/nrendo.2013.58.

23. Bassett JH and Williams GR. Role of Thyroid Hormones in Skeletal Development and Bone Maintenance. Endocr Rev 2016; 37: 135–187. 2016/02/11. DOI: 10.1210/er.2015-1106.

24. Monfoulet LE, Rabier B, Dacquin R, et al. Thyroid hormone receptor beta mediates thyroid hormone effects on bone remodeling and bone mass. Journal of bone and mineral research : the official journal of the American Society for Bone and Mineral Research 2011; 26: 2036–2044. 2011/05/20. DOI: 10.1002/jbmr.432.

25. Wang X, Levy Y and Iwahara J. Competition between Nucleic Acids and Intrinsically Disordered Regions within Proteins. Acc Chem Res 2025 20250709. DOI: 10.1021/acs.accounts.5c00261.

